# Built environment characteristics and drowning mortality: A global satellite-based analysis of urbanisation, infrastructure, and water proximity

**DOI:** 10.64898/2026.04.19.26351236

**Authors:** Ryan Essex, Samsung Lim, Jagnoor Jagnoor

## Abstract

Drowning remains a major global public health challenge, yet how built environment characteristics shape population-level drowning risk remains poorly understood. This study linked satellite-derived built environment data to subnational drowning mortality estimates across 203 regions in 12 countries from 2006–2021. It found that built environment associations with drowning mortality are complex, non-linear, and shaped by development context. Urban extent was strongly protective, while built area near water showed protection overall but increased risk when combined with high population crowding. Almost all drowning mortality variance occurred between regions rather than within regions over time, indicating risk is predominantly determined by place-based characteristics. Income-stratified analyses revealed profound heterogeneity: crowding was protective in low-to middle-income settings but near-null in high-income regions, while waterfront development captured very different realities across contexts. These findings highlight the importance of tailoring drowning prevention strategies to local built environment configurations and development contexts.

## Introduction

Drowning remains a major global public health challenge, responsible for around 300,000 deaths annually (World Health Organization, 2024). While drowning risk factors have traditionally focused on individual behaviours and supervision, the built environment, encompassing urbanisation patterns, infrastructure quality, settlement density, and proximity to water bodies, represents a critical but understudied determinant of population-level drowning mortality.

The literature that exists in this space points to the fact that the built environment matters. Much of this work has focused on differences between rural and urban areas. In a study carried out in Ontario Canada, drowning rates between 2004-2008 were higher in rural areas (Fralick et al., 2013). This pattern persisted in other studies, with drowning rates, amongst other forms of unintentional injury, higher in rural areas in the US (Garnett et al., 2021) and China (Li et al., 2023). Systematic reviews suggest that, like many other risk factors, people in rural areas in low and middle income countries may be particularly vulnerable (Tyler et al., 2017). Elsewhere we find evidence that proximity to water matters. In Bangladesh for example, amongst children, most drowning happened in ponds and ditches, and 80% occurred within 20m of the home, with children in rural areas at highest risk (Rahman et al., 2009). Similar studies suggest that the proximity of water to homes and other infrastructure is a critical factor when it comes to drowning risk (Alam et al., 2025). Moving beyond the simple rural/urban binary, Dai et al. (2013) utilised geographic information system (GIS) data to examine drowning throughout Georgia in the US, finding that amongst higher density drowning neighbourhoods, socioeconomic factors like household income and education levels shaped drowning risk.

While we can piece together a picture about the built environment and exposure to water, the literature in this space is largely in its infancy. The rural-urban divide often drawn in the literature hardly captures the diversity of these environments. While greater urbanisation may be protective in offering better infrastructure, including supervised swimming facilities and physical barriers separating populated areas from water bodies, this may not be the case everywhere. Rapid unplanned urbanisation, for example, may create hazards through informal waterfront settlements, inadequate drainage infrastructure and unsafe water access points. At the same time, while it is well established that factors like age and socioeconomic status influence drowning risk, we know very little about the specific features of how the built environment shapes drowning risk, particularly at neighbourhood and population levels (Scarr & Jagnoor, 2024). Many questions remain unanswered about how built environment characteristics, urbanisation density, infrastructure quality, settlement patterns, or population-water interfaces, shape drowning risk, and whether these associations differ systematically by development context. The need to close this knowledge gap is pressing. The United Nations projects that 68% of the world’s population will live in urban areas by 2050, with 90% of this growth occurring in Asia and Africa (United Nations, 2018). Understanding how built environment characteristics relate to drowning risk has the potential to inform urban planning, infrastructure investment, and targeted prevention strategies.

Fortunately, there is now a substantial body of global, spatially explicit data that can be drawn upon. Satellite-derived data can quantify where water is persistent versus seasonal, allowing measures such as water coverage and distance-to-water to be calculated consistently across settings (Pekel et al., 2016). Global settlement data provides a way to move beyond rural/urban binaries by measuring the texture of human settlement, built-up extent, settlement density, and patterns of development at fine spatial resolutions (Pesaresi et al., 2024). In this study we draw on these data sources to examine associations between built-environment characteristics and drowning mortality at the population level, linking satellite-derived measures to subnational estimates from the Global Burden of Disease (GBD) study.

## Research Aims

This study sought to explore relationships between satellite-derived built environment characteristics and subnational drowning mortality rates globally from 2006 to 2021. Specifically, we sought to firstly quantify associations between urbanisation metrics (urban extent, growth rate, density patterns), infrastructure quality proxies (nighttime lights, informal settlement indicators), settlement patterns (sprawl versus compactness), population crowding, and built environment-water interfaces with age-standardised drowning mortality rates; secondly, test for non-linear relationships, in relation to urban density and development transitions; and thirdly, examine whether built environment effects differ systematically by country income level.

## Methods

### Overview

This study employed a longitudinal ecological design linking satellite-derived built environment variables to subnational drowning mortality rates. Built environment data were extracted from Global Human Settlement Layer (GHSL) (Pesaresi et al., 2024), VIIRS/DMSP nighttime lights (NASA/NOAA; NOAA), WorldPop (WorldPop, 2025) population estimates, and JRC Global Surface Water (Pekel et al., 2016) datasets using Google Earth Engine (GEE) (Gorelick et al., 2017). Analysis focused on subnational regions (ADM1 level) with drowning mortality estimates from the Global Burden of Disease (GBD) 2021 study (Xie et al., 2025) that could be matched with Global Administrative Unit Layer (GAUL) 2015 (Food and Agriculture Organization of the United Nations, 2015) administrative boundaries (n= 203 regions across 12 countries covering 2006-2021). Analysis was conducted in R (version 4.5.0) (Posit team, 2025) using GEE (Gorelick et al., 2017) for data extraction. A full list of all packages used in analysis is included in Table S1 (supplementary material).

### Data extraction and transformation

For each region–year (2006–2021), we used GEE to extract built environment and population characteristics. Subnational boundaries were taken from FAO GAUL (2015) ADM-1 units, with geometries simplified to a 5 km tolerance to reduce computational load. Built-up area was derived from the Global Human Settlement Layer (GHSL P2023A), population from WorldPop, surface water from the JRC Global Surface Water dataset, and infrastructure quality proxies (nighttime lights) from VIIRS (2014 onwards) and DMSP (for earlier years). GHSL and WorldPop data were processed at 100 m resolution, JRC water layers at 30 m (aggregated to 100 m), and nighttime lights at 1 km.

From these sources, we constructed a suite of indicators capturing five domains of the built environment. First, urbanisation level and change were characterised using total built-up area, the share of land that was built-up (“urban extent”). Second, population density and crowding were measured using total population, overall population density, and crowding within built-up areas (population per km^2^ of built surface), including log-transformed and non-linear terms to account for skew and potential threshold effects. Third, waterfront exposure was captured by overlaying built-up areas and population with distance-to-water surfaces from JRC Global Surface Water. We quantified the area of built environment within 100m and 500 m of water, the proportion of total built area near water, population and crowding in waterfront built areas, and the length of the built–water boundary as an indicator of water interface intensity. Finally, infrastructure quality and informality were approximated using mean night-time light intensity and its spatial variation, normalised by built-up area to produce indices of formal infrastructure coverage, informal settlement, and “infrastructure lag” where new development outpaced lighting-based infrastructure.

In total, we extracted or derived over 20 indicators across these domains. Many were used only to construct composite indices or categorical classifications. A summary of the variables included in models is included below (Table 1). Full operational definitions, spatial and temporal resolution, and derivation formulas are provided in Tables S2 (supplementary material). In total, 16 variables were explored as candidates for the main and income-stratified models, with 8 retained. Two variables were used to test non-linear urbanisation effects.

**Table 1.** Summary and description of variables included in modelling.

| Variable | Model | Description |
| --- | --- | --- |
| Urban extent (% built-up area) | LASSO/main and income stratified models; Quadratic model | Proportion of each region classified as built-up; proxy for overall level of urbanisation / settlement density. |
| Urban extent squared (quadratic term) | Quadratic model | Non-linear (quadratic) term for urban extent capturing curvature in the density–drowning relationship (e.g. U-shaped or inverted-U patterns). |
| Annual urbanisation rate (% per year) | LASSO (offered) | Average annual percentage growth in built-up area over the previous ~5 years; proxy for speed of urban expansion. |
| Log crowding in built-up areas | LASSO/main and income stratified models | Log-transformed built-area crowding to reduce skew and allow diminishing marginal effects of very high crowding. |
| Overall population density (persons/km<sup>2</sup>) | LASSO (offered) | Population density including both built-up and non-built areas; broad measure of settlement intensity. |
| Built-up area near water (% of built-up) | LASSO (offered) | Share of built-up area located within 500 m of surface water; proxy for waterfront or waterside development. |
| Crowding in waterfront built areas | LASSO (offered) | Population density in built-up areas close to water; highlights crowded waterfront or informal riverside/coastal settlements. |
| Water interface intensity (km interface/km<sup>2</sup>) | LASSO (offered) | Amount of built–water edge per km<sup>2</sup> of built-up area; distinguishes compact waterfront cores from more dispersed edge development. |
| Sprawl index (settlements per km<sup>2</sup> built) | LASSO (offered) | Degree of fragmentation of built-up areas; higher values indicate more, smaller settlements (i.e. more sprawl) per unit built-up area. |
| Development pattern (sprawl vs compact index) | LASSO/main and income stratified models | Composite index contrasting fragmentation with density; higher values reflect sprawling, low-density development versus compact urban cores. |
| Night lights per unit built area | LASSO/main and income stratified models | Average visible night-time illumination per unit built-up area; proxy for infrastructure and service intensity in developed areas. |
| Night lights coefficient of variation | LASSO/main and income stratified models | Relative spatial inequality in night-time illumination across built-up areas; higher values indicate more uneven infrastructure distribution. |
| Informal settlement index (low lights per built) | LASSO (offered) | Index of areas with relatively dense built-up area but low night-time illumination; proxy for informal or under-served settlements. |
| New development (% of built in last ~10 years) | LASSO (offered) | Share of current built-up area that has appeared in the last ~10 years; indicator of recent or rapid urban expansion. |
| Infrastructure lag in new development | LASSO/main and income stratified models | Extent of new development that remains relatively dark at night; proxy for urban growth outpacing formal infrastructure/electrification. |
| Built-up area (current year; km<sup>2</sup>, 100 m) | LASSO/main and income stratified models | Total built-up area in each region for the analysis year; measure of built extent/scale. |
| Crowding–water interaction | LASSO/main and income stratified models | Relative concentration of crowding in waterfront built areas compared with overall built-up crowding. |

### Analytic strategy

#### Overview

A multi-stage statistical analysis to examine associations between satellite-derived built environment characteristics and drowning mortality rates across 203 sub national regions between 2006–2021 was conducted. Analysis proceeded in six stages. First, exploratory descriptive analyses; Second, variable selection for subsequent modelling; Third, main (mixed-effects) modelling examining associations between built environment variables in drowning rates; Fourth, stratification of main model by income group (low, low-middle, high-middle and high income countries); Fifth, non-linear (quadratic) exploration of the relationship between urbanisation and drowning rates; Sixth, diagnostic, sensitivity and robustness checks. Each step will be expanded upon below.

#### Descriptive statistics

We first characterised the distribution of drowning mortality rates and built environment variables across regions and time periods. For continuous variables, we calculated means, standard deviations, medians, interquartile ranges, and ranges. We examined variable distributions to inform modelling decisions and assessed temporal trends. To understand heterogeneity across development contexts, we calculated stratified descriptive statistics by World Bank income group (low income, lower-middle income, upper-middle income, and high income, using 2021 classifications).

#### Variable selection

Given the large number of potential built environment predictors (n = 16) and concerns about multicollinearity among urbanisation measures, we conducted preliminary correlation and univariate analyses to identify the most relevant variables. Based on these analyses and theoretical considerations about drowning mechanisms, we selected nine variables to enter into the Least Absolute Shrinkage and Selection Operator (LASSO) regression: crowd-water interaction, crowding (log-transformed), urban extent, built area within 100m of water, infrastructure lag, development pattern (sprawl vs compact), nighttime lights inequality (coefficient of variation), infrastructure quality (nighttime lights per built area), and informal settlement index.

LASSO applies L1 regularisation to shrink coefficients of less important predictors toward zero, effectively performing variable selection while reducing overfitting. All continuous predictors were standardised (mean = 0, standard deviation = 1) prior to LASSO to ensure comparability of penalisation across variables measured on different scales. We used 10-fold cross-validation to select the regularisation parameter (λ), choosing λ.min (to minimise cross-validation error) to maximise predictive performance. Variables retained by LASSO (non-zero coefficients) were carried forward to the main mixed-effects models.

To verify stability of variable selection, we conducted sensitivity analyses testing. First, alternative regularization parameters (λ.min vs λ.1se, where λ.1se represents the most parsimonious model within one standard error); Second, elastic net regularisation (α = 0.5) versus pure LASSO (α = 1.0), and; Third, consistency across five different random seeds for cross-validation folds. Variable selection showed strong stability, with >80% overlap across alternative specifications.

#### Main Mixed-Effects Model

Our primary analytical model was a linear mixed-effects regression with random intercepts by region to account for the nested structure of annual observations within subnational regions. The model took the form:

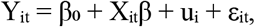

where Y_it_ was the age-standardized drowning rate per 100,000 for region *i* in year *t*, X_it_ was a vector of standardized built environment predictors, β was a vector of fixed effect coefficients, u _i_ was a random intercept for region i (assumed normally distributed with mean 0 and variance σ _u_^2^), and ε_it_ was the residual error term (assumed normally distributed with mean 0 and variance σ_e_^2^). The model family was Gaussian and was fit using restricted maximum likelihood (REML) to obtain unbiased variance estimates.

#### Income Stratification

To test whether built environment effects varied by development context, we stratified analyses by World Bank (2021) income group. We fit separate mixed-effects models (identical structure to the main model) within each income stratum. This approach allowed effect sizes to vary freely across income groups rather than constraining them through interaction terms.

#### Non-linear urbanisation effects

We tested for non-linear relationships between urban extent and drowning mortality by fitting a quadratic ordinary least squares regression model:

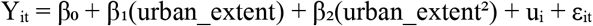

This was a simple regression model without random effects, as it served as an exploratory analysis of the functional form of the urbanization-drowning relationship. We calculated the critical point (-β_1_/2β_2_) to identify the urban extent level at which drowning risk was minimized or maximized. A positive β_2_ coefficient indicated a U-shaped relationship (risk declining then increasing with urbanisation), while negative β_2_indicated an inverted-U. We compared quadratic versus linear models to determine whether the quadratic term significantly improved model fit.

#### Diagnostics and Robustness

We conducted comprehensive diagnostics to verify model assumptions. For the outcome, we assessed normality using Shapiro–Wilk tests and summarised distributional characteristics (including skewness and proportion of zeros). For residuals, we examined Pearson residual summaries and undertook model checking using DHARMa simulated residuals with tests for uniformity, dispersion, and outliers (with diagnostic plots saved). We justified random effects by comparing information criteria across fixed-effects (no random effects), random-intercepts, and (where convergence permitted) random-slopes specifications, and calculated intraclass correlation coefficients (ICC). We assessed multicollinearity via variance inflation factors (with VIF<5 treated as acceptable and VIF<10 as moderate), after checking for aliased coefficients due to perfect collinearity.

We also conducted seven robustness checks. First, alternative outcome specifications (Gaussian vs Gamma with log link where feasible vs log-transformed Gaussian); second, random effects structure (no random effects vs random intercepts vs random slopes for key predictors); third, influential observations (Cook’s distance screening; refitting after excluding influential observations when the number was small, and flagging >20% coefficient change); fourth, sample restrictions (excluding regions with <10 years of data, winsorizing outcomes at the 1st/99th percentiles, and excluding low-income countries where income-group classification was available); fifth, LASSO sensitivity (λ.min, elastic net, and varying random seeds); sixth, bootstrap inference (200 resamples to compare bootstrap vs model-based standard errors and confidence intervals), and; seventh, variable transformations (alternative specification of crowding—linear vs log—where applicable, and a square-root transformation of the outcome).

## Results

### Descriptive statistics

The analytic sample comprised 3,248 region-year observations across 203 subnational regions (2006–2021). The outcome (drowning mortality rate per 100,000) had a mean of 2.64 (SD 1.87), with median 2.28 (IQR 1.18–3.47) and range 0.23–11.01. The sample included 7 low-income regions (112 observations from Ethiopia), 29 lower-middle income regions (464 observations from India, Kenya, and Pakistan), 76 upper-middle income regions (1,216 observations from Brazil, Iran, Mexico, and South Africa), and 91 high-income regions (1,456 observations from Italy, Japan, Norway, and the United States). All 203 regions contributed 16 years of data. Built environment measures varied substantially across settings: urban extent averaged 1.68% (SD 2.95; median 0.71%), log population crowding averaged 8.82 (SD 0.72; median 8.77), and built area within 100m of water averaged 6.29 km^2^ (SD 10.23; median 2.41 km^2^). The infrastructure lag index had mean 10.86 (SD 7.37; median 8.70), and nighttime light metrics showed wide dispersion across regions (coefficient of variation: mean 0.69, SD 0.61; light-per-built-area: mean 26,679, SD 452,977), indicating substantial heterogeneity in the intensity and regularity of development. All descriptive statistics are included in Tables S3-5 and Figure S1 (supplementary material).

### Variable selection

To identify variables for multivariate modelling, we prioritised rates, ratios, and indices rather than absolute measures, to enable fairer comparison across regions of very different sizes and baseline development levels. From the assembled data we considered 16 built environment measures spanning urbanisation (urban extent), density/crowding (including log crowding), water proximity (including built area within 100m of water), settlement morphology (development pattern, sprawl), and infrastructure proxies (nighttime lights and infrastructure lag), alongside interaction terms capturing the coupling of crowding and water exposure.

Bivariate correlations between drowning rates and built environment predictors, and correlations among predictors (to assess multicollinearity), are reported in Tables S6 and S7 (supplementary material). Univariate OLS models (each regressing drowning rate on a single standardised predictor) were broadly consistent with the bivariate patterns, though effect directions for several predictors reversed after adjustment (Table S8). The largest univariate association was observed for the crowding × water interaction (β = 0.87 per SD increase; R^2^ = 0.22), consistent with drowning risk clustering where dense settlement coincides with high water exposure. Univariate associations were also positive for infrastructure lag (β = 0.58; R^2^= 0.10) and log crowding (β = 0.53; R^2^ = 0.08), while urban extent (β = −0.19) and built area within 100m of water (β = −0.32) showed protective univariate associations despite explaining relatively little variance individually.

From these 16 potential predictors, we refined the set to 9 variables based on bivariate associations, conceptual relevance, and collinearity screening (retaining one variable from each highly correlated pair). This pre-selection reduced dimensionality while preserving distinct aspects of the built environment. We then entered 9 variables into LASSO regression. We used 10-fold cross-validation to select the regularisation parameter (λ) and applied the λ.min criterion to minimize cross-validation error. LASSO retained eight predictors spanning crowding (log crowding), water proximity (built area within 100m of water), urbanisation (urban extent), infrastructure proxies (nighttime light per built area; nighttime light coefficient of variation; infrastructure lag), settlement morphology (development pattern), and the crowding × water interaction. Coefficient signs in LASSO aligned with the direction of associations in the fully adjusted mixed-effects models described below (noting that several differed from the univariate associations), reinforcing that the strongest signals emerged only when accounting for the joint structure of development, density, and water exposure. These results are included in Table S9 (supplementary material).

### Main mixed-effects model

The mixed-effects model with random intercepts by region (N = 3,248 observations from 203 regions), using the eight LASSO-selected predictors, revealed several consistent associations.

Higher urban extent remained strongly protective (β = −0.80, SE = 0.11, p < 0.001), indicating that, after adjustment for other built environment characteristics, more urbanised regions tended to have lower drowning mortality. Log population crowding showed a similarly strong protective association (β = −1.34, SE = 0.07, p < 0.001).

Water-proximity measures and their coupling with crowding showed a more nuanced pattern. Built area within 100m of water was strongly protective (β = −0.88, SE = 0.07, p < 0.001), while the crowding × water interaction was positively associated with drowning (β = 0.25, SE = 0.06, p < 0.001). Taken together, these results suggest that regions with more built development close to water tend to have lower drowning rates (potentially reflecting more managed waterfronts and associated infrastructure), but risk rises where water exposure coincides with high crowding, consistent with a concentration of exposure in dense communities with direct water access.

Settlement morphology was also independently associated with risk. The development pattern index showed a protective association (β = −0.14, SE = 0.03, p < 0.001), suggesting that more coherent, less fragmented development patterns are associated with lower drowning mortality, even after accounting for density and urban extent.

Infrastructure proxies were statistically significant but comparatively smaller in magnitude. Infrastructure lag was protective (β = −0.31, SE = 0.02, p < 0.001). Nighttime light coefficient of variation was also protective (β = −0.10, SE = 0.01, p < 0.001). Nighttime light per built area was small and non-significant (β = 0.02, SE = 0.01, p = 0.205). Model fit statistics indicated adequate performance (AIC = 4,260.50; BIC = 4,327.44). The intraclass correlation coefficient (ICC = 0.980) indicated that 98.0% of total variance in drowning rates occurred between regions, with only 2.0% occurring within regions over time, reinforcing that drowning mortality in this dataset is predominantly differentiated cross-sectionally rather than temporally within regions. These results are summarised in Figure 1 and Table S10 (supplementary material).

**Figure 1.**
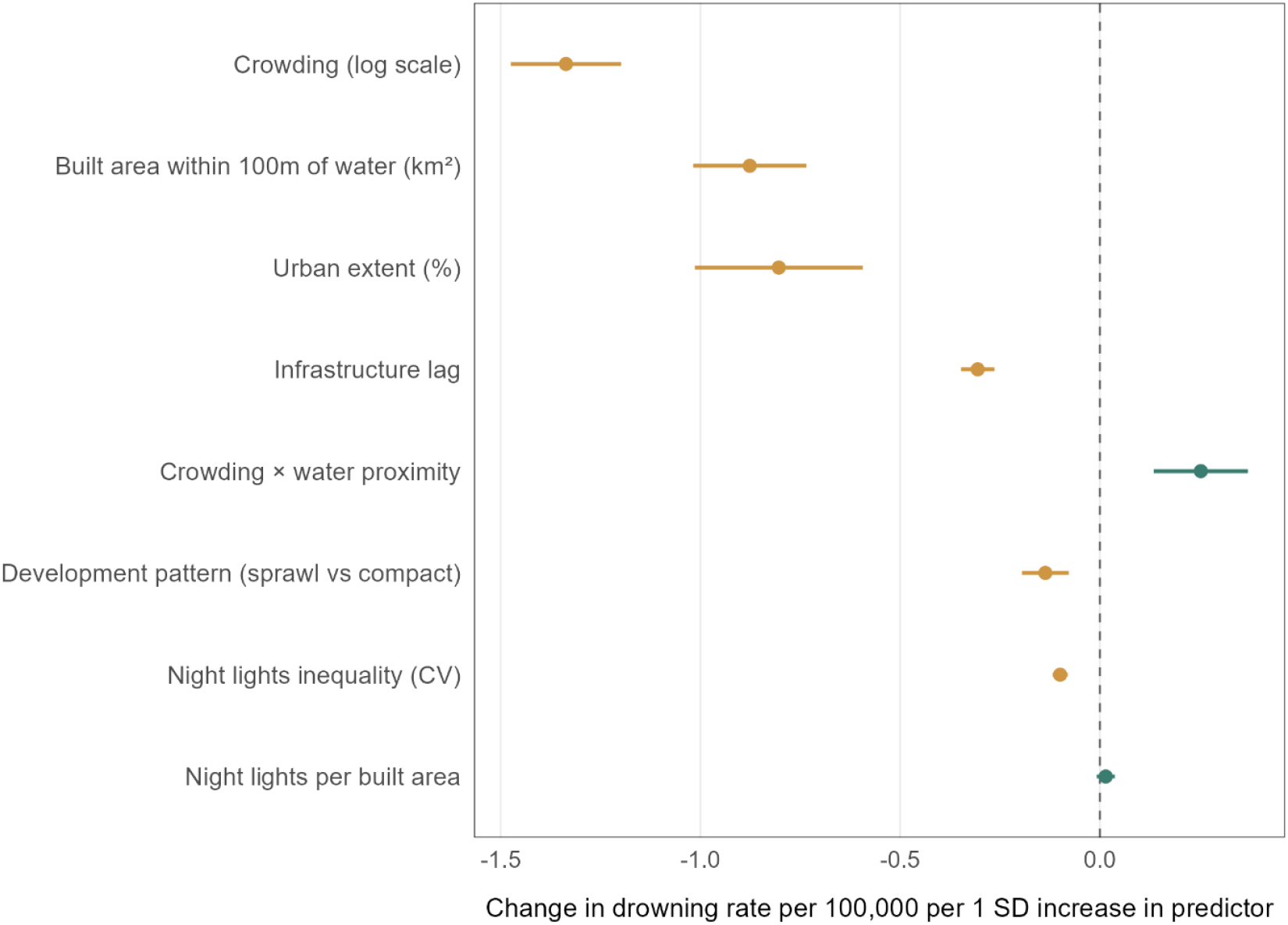
Main model: Built environment predictors of drowning mortality

### Income stratification

Stratified analyses revealed substantial heterogeneity in built environment associations across income levels. The low-income stratum (n = 112 observations from 7 regions in 1 country) showed unstable coefficient magnitudes for some predictors (notably built area within 100m of water), reflecting limited sample size and likely context-specific heterogeneity; these results should be interpreted with caution.

Several associations were broadly consistent across settings. Urban extent remained protective in all income strata (β from −0.24 in high-income to −1.46 in upper-middle-income; all p < 0.001). Infrastructure lag was protective in low-, lower-middle-, and upper-middle-income strata but was positive in the high-income stratum, suggesting that the meaning of “lag” may differ in more developed contexts (or may proxy different mechanisms once baseline infrastructure is high).

Most notably, the crowding × water interaction differed sharply by context. It was positively associated with drowning in low-income regions (β = 0.45, p < 0.001), near-null in lower-middle-income regions (β = 0.05, p = 0.325) and became protective in upper-middle- and high-income regions (β = −0.53, p < 0.001; and β = −0.16, p = 0.024, respectively). This inversion suggests that the same “density × water exposure” configuration may imply very different risk environments depending on the maturity of infrastructure, formal regulation of water access, and emergency response capacity.

Built area within 100m of water similarly differed by income group: strongly protective in lower-middle- and upper-middle-income strata (β = −2.45 and −10.11, both p < 0.001), but non-significant in high-income settings and highly unstable in the low-income stratum. Nighttime light coefficient of variation was generally protective outside the low-income stratum. Nighttime light per built area was non-significant in low- and lower-middle-income strata, strongly positive in upper-middle-income regions, and near-zero but statistically significant in high-income regions, again highlighting that proxy indicators for “infrastructure” or “development intensity” behaved differently across development contexts. These findings are summarised in Figure 2 and Table S11 (supplementary material).

**Figure 2.**
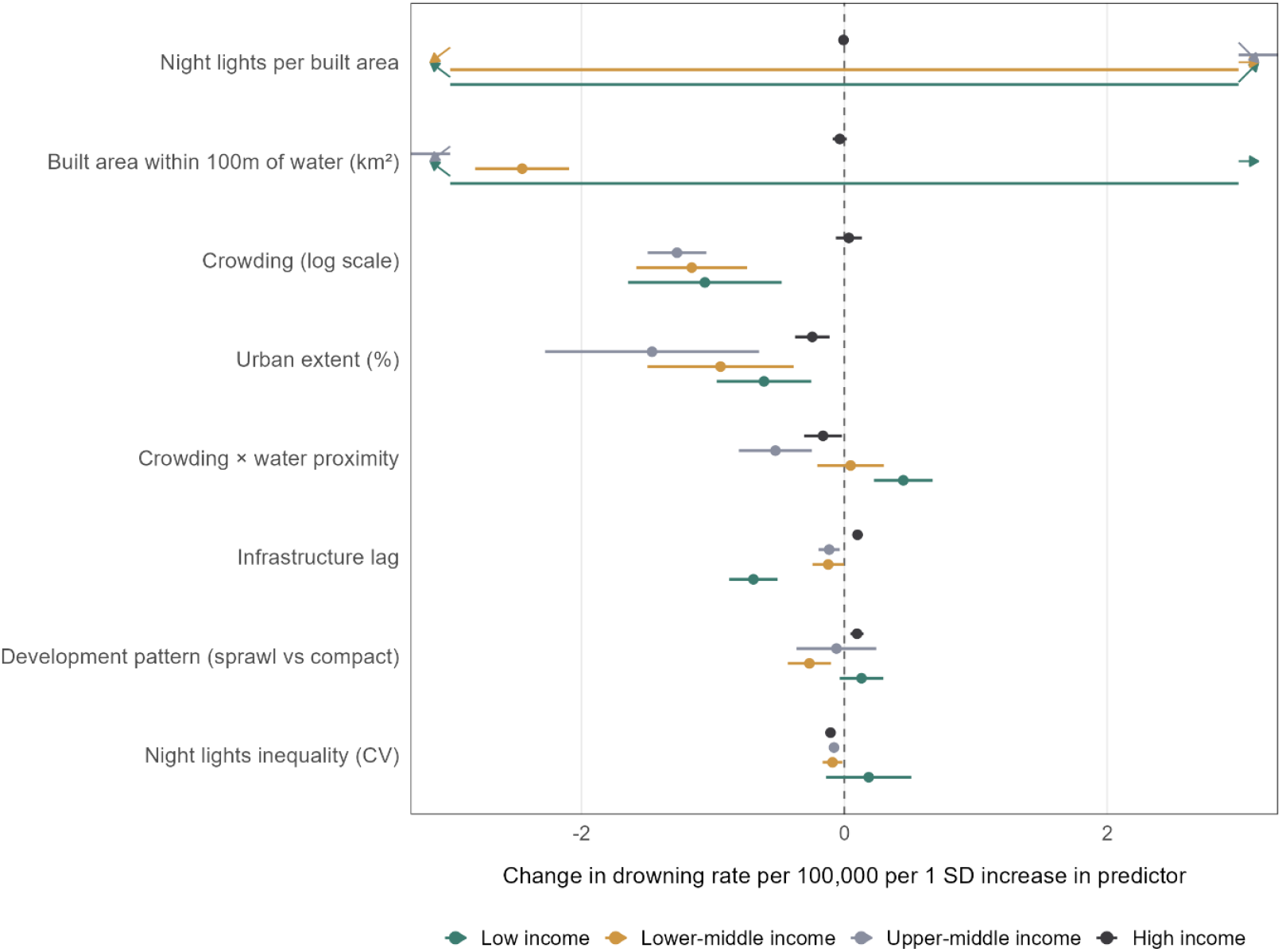
Income stratified model

### Non-linear urbanisation effects

To test for non-linear relationships, we utilised quadratic models including both linear and squared terms for urban extent. The quadratic model confirmed substantial non-linearity: the linear term for urban extent was negative (β = −0.279, p < 0.001), while the squared term was positive (β = +0.014, p < 0.001), consistent with a U-shaped relationship. This pattern suggested that drowning mortality initially declined with urbanisation from very low levels, reached a minimum at intermediate urbanisation (critical point ~9.9% urban extent), then increased at higher urbanisation levels. As above, this aggregate curve should be interpreted alongside the income-stratified results, which show that the mechanisms linking urbanisation to drowning differ substantially by development context. These findings are summarised in Figure 3 and Table S12 (supplementary material).

**Figure 3:**
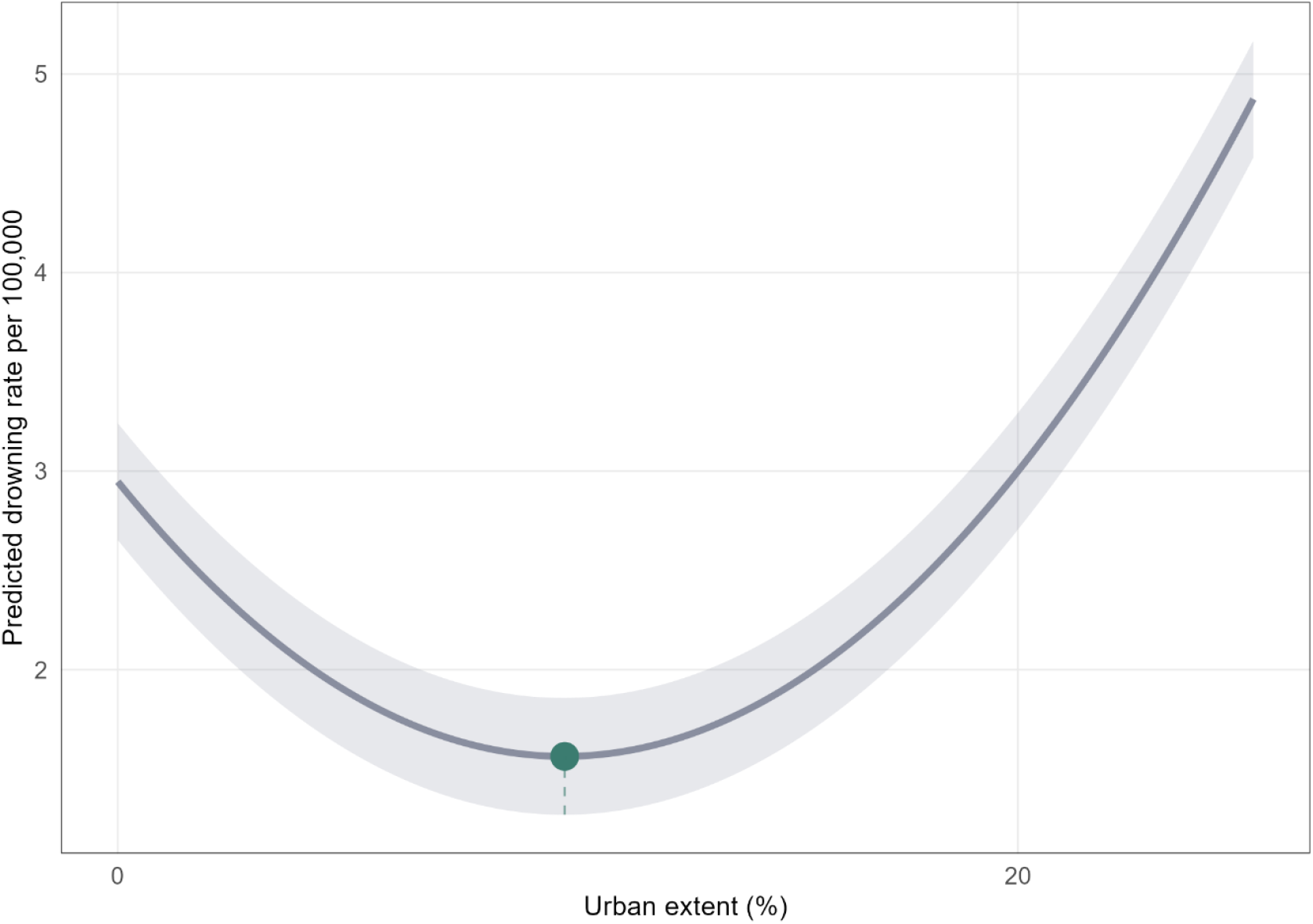
Quadratic model: Non-linear relationship between urban extent and drowning mortality

### Model diagnostics

Diagnostic checks informed our final specification. The outcome distribution (mean = 2.64 per 100,000; SD = 1.87) showed modest skew and minimal zeros, supporting a Gaussian mixed modelling approach for interpretability. Random effects structure analyses strongly favoured mixed-effects models over fixed-effects-only specifications (AIC 4,260.50 vs 12,176.78), reflecting substantial between-region heterogeneity. Random slopes provided markedly better fit in aggregate but were not feasible across the full stratified and temporal suite of models, and we therefore retained random intercepts for consistent inference across analyses. The ICC of 0.980 indicates that most variation occurs between regions, justifying the random intercept approach to account for persistent, unmeasured regional characteristics (e.g., enforcement, water safety infrastructure quality, and emergency response capacity) not captured by measured predictors. Multicollinearity among the eight retained predictors was low (VIF range approximately 1.15–1.84), consistent with LASSO having reduced redundancy while preserving key dimensions of the built environment. These findings are summarised in Tables S13 (supplementary material).

### Sensitivity and robustness

We assessed whether the main findings were sensitive to modelling assumptions, influential observations, sample restrictions, and variable-selection choices. Alternative distributional assumptions and outcome transformations were evaluated as sensitivity checks; while some alternatives improved statistical fit, the substantive conclusions were unchanged, with urban extent and established development remaining inversely associated with drowning risk and the crowding–water interaction remaining harmful. We examined random-effects structure by comparing models with no random effects, random intercepts, and random slopes for urban extent; allowing random slopes improved fit, indicating heterogeneity in the urbanisation– drowning relationship across regions, but the main fixed-effect conclusions were consistent. Additionally, we retained random intercepts as the primary specification because the main objective was inference on average associations rather than region-specific slopes and random-slopes models increase complexity and can yield unstable variance–covariance estimates in settings with limited within-region variation. We evaluated influential observations using Cook’s distance; >5% of observations exceeded conventional thresholds, suggesting heterogeneity is a structural feature of the data rather than being driven by a small number of outliers, and therefore exclusion was not pursued. Sample sensitivity checks (including winsorisation of extreme outcome values) did not alter the direction or statistical support of key coefficients. Finally, LASSO selection was robust to alternative penalisation choices: λ.min selected more predictors than λ.1se, but retained all predictors from the main model, indicating stable identification of the core built-environment signals. Bootstrap resampling provided an additional check on uncertainty estimates; while bootstrap-to-model SE ratios suggested modest discrepancies for some parameters, confidence intervals were substantively consistent. These findings are summarised in Tables S14(supplementary material).

## Discussion

This study leveraged satellite-derived built environment data linked to subnational drowning mortality across 203 regions in 12 countries from 2006–2021 to examine how urbanisation characteristics relate to drowning risk. We found that built environment associations with drowning mortality are complex, non-linear, and shaped by development context. Overall, more established urbanisation was associated with lower drowning risk, but the relationships depended critically on how populations and development were distributed around water. Critically, almost all drowning mortality variance (~98.0%) occurred between regions rather than within regions over time, indicating drowning risk was predominantly determined by place-based characteristics that remain relatively stable.

Urban extent showed a strong protective association (β = −0.804, p < 0.001), consistent with formal development reducing drowning through infrastructure provision and reduced incidental and occupational exposure to water, amongst other mechanisms. However, several results indicate that “development near water” was not uniformly protective or harmful; rather, risk depended on how development and population concentrate at the water’s edge. Greater built area within 100m of water (km^2^) was strongly protective overall (β = −0.876, p < 0.001), while the crowding × water interaction term was positive (β = +0.253, p < 0.001), indicating that risk increases where high crowding co-occurs with substantial near-water built environments. This suggests that waterfront development may reduce risk when it reflects managed access and mature infrastructure, for example, but that the combination of concentrated populations and near-water development can create elevated risk where exposure and infrastructure strain coincide.

Crowding was also influential. Log population crowding was strongly protective overall (β = −1.336, p < 0.001). But this pattern varied sharply by income group: crowding remained strongly protective across low-, lower-middle-, and upper-middle-income strata, yet was near-null in high-income regions. This heterogeneity implies that in actively urbanising settings, crowding may proxy for formal, serviced settlement structures, whereas in high-income settings crowding may be less informative because baseline safety infrastructure is more uniform. The development pattern index was protective overall (β = −0.136, p < 0.001), but its stratified effects were not uniform (protective in lower-middle income; risk in high income), reinforcing that “urban form” operates differently depending on governance, baseline infrastructure, and where growth is occurring.

Income-stratified analyses confirmed profound heterogeneity in how built environment features relate to drowning. Urban extent was protective in every income stratum (β = −0.612 to −1.464 in low to upper-middle income; β = −0.244 in high income; all p < 0.001). The near-water built area term showed strong protection in lower-middle and upper-middle income settings, but was non-significant in high income and imprecise in low income, consistent with waterfront development capturing very different realities across contexts. The crowding × water interaction also differed by development context (positive in low income; negative in upper-middle and high income), suggesting that the same physical configuration, dense settlement near water, may signal unmanaged exposure in some settings and managed, regulated environments in others.

Overall, these results add vital context and nuance to the existing literature. Our overall finding, that infrastructure and urbanisation can act to lessen drowning risk, is already well supported (Tan et al., 2023). Our finding that most variance existed between regions can also be found throughout the existing literature on injury prevention, where structural, place-based determinants have been found to be influential in their association to risk of injury (Hameed et al., 2010; McClure et al., 2015). Perhaps most importantly however, these findings add nuance to the longstanding recognition that when it comes to injury prevention, our environment matters and can play a critical role in determining exposure and risk when it comes to drowning (Stevenson, 2006).

### Limitations

This study has several limitations. First, the analysis is ecological and observational, and we cannot make causal claims about built environment characteristics and drowning mortality. Residual confounding is likely. Second, while satellite-derived built environment indicators provide consistent cross-national measurement, they remain proxies. For example, “infrastructure lag” and nighttime light metrics capture broad development intensity and inequality but cannot directly measure drainage systems, barriers, safe water access points, or enforcement of building codes. Likewise, the “built area within 100m of water” measure does not distinguish safe, managed waterfronts from hazardous informal settlements or unprotected riverbanks. Third, the very high intraclass correlation indicates that most variation occurs between regions; consequently, within-region signals are comparatively weak. Finally, income-stratified analyses should be interpreted cautiously at the extremes. The low-income stratum is small (one country, seven regions), limiting precision and generalisability. Conversely, in high-income strata, reduced heterogeneity in infrastructure and safety systems may make built-environment proxies less informative, even if local environments remain important.

## Conclusions

Despite these limitations, these findings have several implications for drowning prevention. First, the results reinforce that drowning risk is strongly place-based: persistent differences between regions far exceed year-to-year changes. This supports targeting strategies that prioritise high-burden regions and invest in durable, structural interventions rather than relying solely on short-term behaviour change programs. Second, the pattern of associations around water exposure suggests a useful distinction between waterfront development and waterfront crowding. Greater near-water built area appears protective on average, likely reflecting managed access and infrastructure, but risk increases where dense populations and near-water development coincide. Practically, this points to interventions that focus on the interface where people and water meet in crowded settings; safe access points, barriers and fencing, drainage and flood management, signage, and rapid response, rather than treating “proximity to water” as uniformly hazardous. Third, the striking heterogeneity by income level suggests that drowning prevention strategies should be tailored to development context. In lower- and middle-income settings, where signals are strongest, integrating drowning prevention into urban planning, housing policy, and informal settlement upgrading may yield the largest gains. Finally, the approach demonstrates the potential value of satellite-derived built environment data for drowning surveillance and prospective risk mapping. These measures can be updated regularly, applied across countries with consistent definitions, and used to identify regions where development and exposure are converging. Future work should link these region-level signals to finer-scale data, subregional drowning events and location-specific hazards, to strengthen causal inference and to translate these findings into operational planning tools for prevention.

## Data Availability

Data used in this study is publicly available

## Notes

### Competing Interest Statement

The authors have declared no competing interest.

### Funding Statement

No funding was received for this study

## References

Alam, E., Al Hattawi, K. S., Akter, H., Alam, J., Alvarez, E., Sufi, F., Islam, M. K., & Islam, A. R. M. T. (2025). Socioeconomic, demographic and environmental factors of child drownings in Northern Bangladesh. Injury Prevention. Advance online publication. doi:10.1136/ip-2024-045434

Dai, D. J., Zhang, Y. Z., Lynch, C. A., Miller, T., & Shakir, M. (2013). Childhood drowning in Georgia: A geographic information system analysis. Applied Geography, 37, 11–22. 10.1016/j.apgeog.2012.10.006

Food and Agriculture Organization of the United Nations. (2015). Global Administrative Unit Layers (GAUL). Food and Agriculture Organization of the United Nations.https://developers.google.com/earth-engine/datasets/catalog/FAO_GAUL_2015_level0

Fralick, M., Gallinger, Z. R., & Hwang, S. W. (2013). Differences in drowning rates between rural and non-rural residents of Ontario, Canada. International Journal of Aquatic Research and Education, 7(4), 6.

Garnett, M. F., Spencer, M. R., & Hedegaard, H. (2021). Urban-rural differences in unintentional injury death rates among children aged 0-17 years: United States, 2018-2019. NCHS data brief, 421, 1–8.

Gorelick, N., Hancher, M., Dixon, M., Ilyushchenko, S., Thau, D., & Moore, R. (2017). Google Earth Engine: Planetary-scale geospatial analysis for everyone. Remote sensing of Environment, 202, 18–27.

Hameed, S. M., Bell, N., & Schuurman, N. (2010). Analyzing the effects of place on injury: Does the choice of geographic scale and zone matter? Open Medicine, 4(4), e171.

Li, Z., Deng, X., Jin, Y., Duan, L., & Ye, P. (2023). Unintentional Drowning Mortality Among Individuals Under Age 20—China, 2013–2021. China CDC Weekly, 5(47), 1058.

McClure, R., Kegler, S., Davey, T., & Clay, F. (2015). Contextual determinants of childhood injury: a systematic review of studies with multilevel analytic methods. American Journal of Public Health, 105(12), e37–e43.

NASA/NOAA. Visible Infrared Imaging Radiometer Suite (VIIRS) Day/Night Band (DNB) data. https://earthdata.nasa.gov/.

NOAA. DMSP OLS: Nighttime Lights Time Series. https://www.ncei.noaa.gov/products/dmsp-operational-linescan-system

Pekel, J.-F., Cottam, A., Gorelick, N., & Belward, A. S. (2016). High-resolution mapping of global surface water and its long-term changes. Nature, 540(7633), 418–422.

Pesaresi, M., Schiavina, M., Politis, P., Freire, S., Krasnodębska, K., Uhl, J. H., Carioli, A., Corbane, C., Dijkstra, L., & Florio, P. (2024). Advances on the Global Human Settlement Layer by joint assessment of Earth Observation and population survey data. International Journal of Digital Earth, 17(1), 2390454.

Posit team. (2025). RStudio: Integrated Development Environment for R. Posit Software, PBC, Boston, MA. http://www.posit.co/

Rahman, A., Mashreky, S. R., Chowdhury, S., Giashuddin, M., Uhaa, I., Shafinaz, S., Hossain, M., Linnan, M., & Rahman, F. (2009). Analysis of the childhood fatal drowning situation in Bangladesh: exploring prevention measures for low-income countries. Injury Prevention, 15(2), 75–79.

Scarr, J.-P., & Jagnoor, J. (2024). Conceptual definition for drowning prevention: A Delphi study. Injury Prevention, 30(2), 145–152.

Stevenson, M. (2006). Building safer environments: injury, safety, and our surroundings. Injury Prevention, 12(1), 1–3.

Tan, H., Lin, Z., Fu, D., Dong, X., Zhu, S., Huang, Z., Liu, Y., He, G., Yang, P., & Liu, T. (2023). Change in global burden of unintentional drowning from 1990 to 2019 and its association with social determinants of health: findings from the Global Burden of Disease Study 2019. BMJ Open, 13(4), e070772.

Tyler, M. D., Richards, D. B., Reske-Nielsen, C., Saghafi, O., Morse, E. A., Carey, R., & Jacquet, G. A. (2017). The epidemiology of drowning in low-and middle-income countries: a systematic review. BMC public health, 17(1), 413.

United Nations. (2018). 68% of the world population projected to live in urban areas by 2050, says UN. https://www.un.org/development/desa/en/news/population/2018-revision-of-world-urbanization-prospects.html

World Health Organization. (2024). Global status report on drowning prevention 2024. World Health Organization.

WorldPop. (2025). WorldPop: Open Spatial Demographic Data and Research. http://www.worldpop.org

Xie, Z., Huang, Z., Ran, Q., Luo, W., & Du, W. (2025). Global burden of drowning and risk factors across 204 countries from 1990 to 2021. Scientific Reports, 15(1), 10916.

